# Gut microbial features and dietary fiber intake predict gut microbiota response to resistant starch supplementation

**DOI:** 10.1101/2023.03.24.23287665

**Authors:** Sri Lakshmi Sravani Devarakonda, Dorothy K. Superdock, Jennifer Ren, Lynn M. Johnson, Aura (Alex) P. Loinard-González, Angela C. Poole

## Abstract

Resistant starch (RS) consumption can have beneficial effects on human health, but the response, in terms of effects on the gut microbiota and host physiology, varies between individuals. Factors predicting the response to RS are not yet established and would be useful for developing precision nutrition approaches that maximize the benefits of dietary fiber intake. We sought to identify predictors of gut microbiota response to RS supplementation. We enrolled 76 healthy adults into a seven-week crossover study. Participants consumed RS type 2 (RS2), RS type 4 (RS4), and a digestible starch, for ten days each with five-day washout periods in between. We collected fecal and saliva samples and food records before and during each treatment period. We performed 16S rRNA gene sequencing and measured fecal short-chain fatty acids (SCFAs), salivary amylase gene copy number, and salivary amylase activity (SAA). Dietary fiber intake was predictive of relative abundance of several amplicon sequence variants (ASVs) at the end of both RS treatments. Treatment order (the order of consumption of RS2 and RS4), alpha diversity, and a subset of ASVs were predictive of SCFA changes after RS supplementation. SAA was only predictive of the relative abundance of ASVs after digestible starch supplementation. Based on our findings, dietary fiber intake and gut microbiome composition would be informative if assessed prior to recommending RS supplementation. Using a precision nutrition approach to optimize the benefits of dietary fibers such as RS could be an effective strategy to compensate for the low consumption of dietary fiber nationwide.

## Introduction

Resistant starch (RS) resists degradation by human enzymes and reaches the large intestine where it can be fermented by colonic microbes to produce beneficial metabolites.^1^ RS is classified into five subtypes (RS1-RS5), and most human studies use RS types 2, 3 and 4. Different microbes can selectively ferment different types of RS to produce short-chain fatty acids (SCFAs).^2^ Butyrate and propionate, two such SCFAs, have numerous health benefits including inducing satiety and improving glucose homeostasis and lipid metabolism.^1,3–5^

Interestingly, several groups have reported interindividual variability in gut microbiota response and host physiological response (e.g. improvements in glucose metabolism) to supplementation with RS2 and RS4.^6–13^ Host and microbial factors prior to RS supplementation may be predictive of, and responsible for the interindividual variability in, gut microbiota response to RS. Previous studies have shown that habitual dietary fiber intake,^14,15^ and baseline microbiome composition^16^ affect gut microbiota response to several nonstarch polysaccharides. Also, host dietary practice can alter microbial diversity, which in turn can affect response to dietary interventions.^17^

In addition, the genotype of the host may influence microbial response. A proposed genetic factor that may predict variable response to RS is *AMY1*, a gene copy number (CN) variant, which encodes the salivary amylase enzyme.^10,18^ *AMY1* CN is correlated with salivary amylase activity (SAA), which helps to initiate starch digestion. *AMY1* CN has been associated with BMI, glucose metabolism, and gut microbiome composition.^18–23^ For this reason, we posited that *AMY1* CN or SAA may be predictive of response to dietary fiber intake. Although differences in preintervention host and microbial characteristics are thought to be responsible for the reported variability, as of yet, the field has not established predictors of gut microbiota or host physiological response to RS supplementation. This knowledge would aid in developing precision nutrition approaches and, in particular, tailoring dietary fiber recommendations to individuals seeking health benefits from RS consumption.

Here, we address a gap in knowledge regarding the factors underlying interindividual variation in gut microbiota response to RS supplementation. In this dietary intervention study, we supplemented participants’ diets with two types of RS, RS2 and RS4, as well as a control digestible corn starch to assess the resulting changes in gut microbiota composition and function. Then we evaluated the ability of the candidate predictors—*AMY1* CN, SAA, prior gut microbiome composition, and dietary fiber intake—to explain interindividual differences in the response to each treatment. Further, because we used a crossover design, we were able to evaluate the effect of the order of treatments on the observed results.

## Results

### Baseline characteristics of study participants

We enrolled 76 individuals into a seven-week open-label crossover dietary intervention study with 59 individuals completing the full study (Figure 1A). Study enrollment and the number of participants completing each treatment are detailed in Figure 1B. We describe baseline characteristics for participants included in our analyses in Table 1. When comparing the two different treatment order groups, Group A and Group B, at baseline, there were no significant differences in age, sex, dietary fiber intake, body fat percent in males, or body fat percent in females (p>0.05 for all). Of note, the average amount of dietary fiber consumed by all participants during the baseline period (11.88 g, SD=4.98 g) was lower than the Institute of Medicine’s recommendations of 25–38 g/day depending on sex and age,^24^ and below the dietary reference intake.^25^

**Figure 1.**
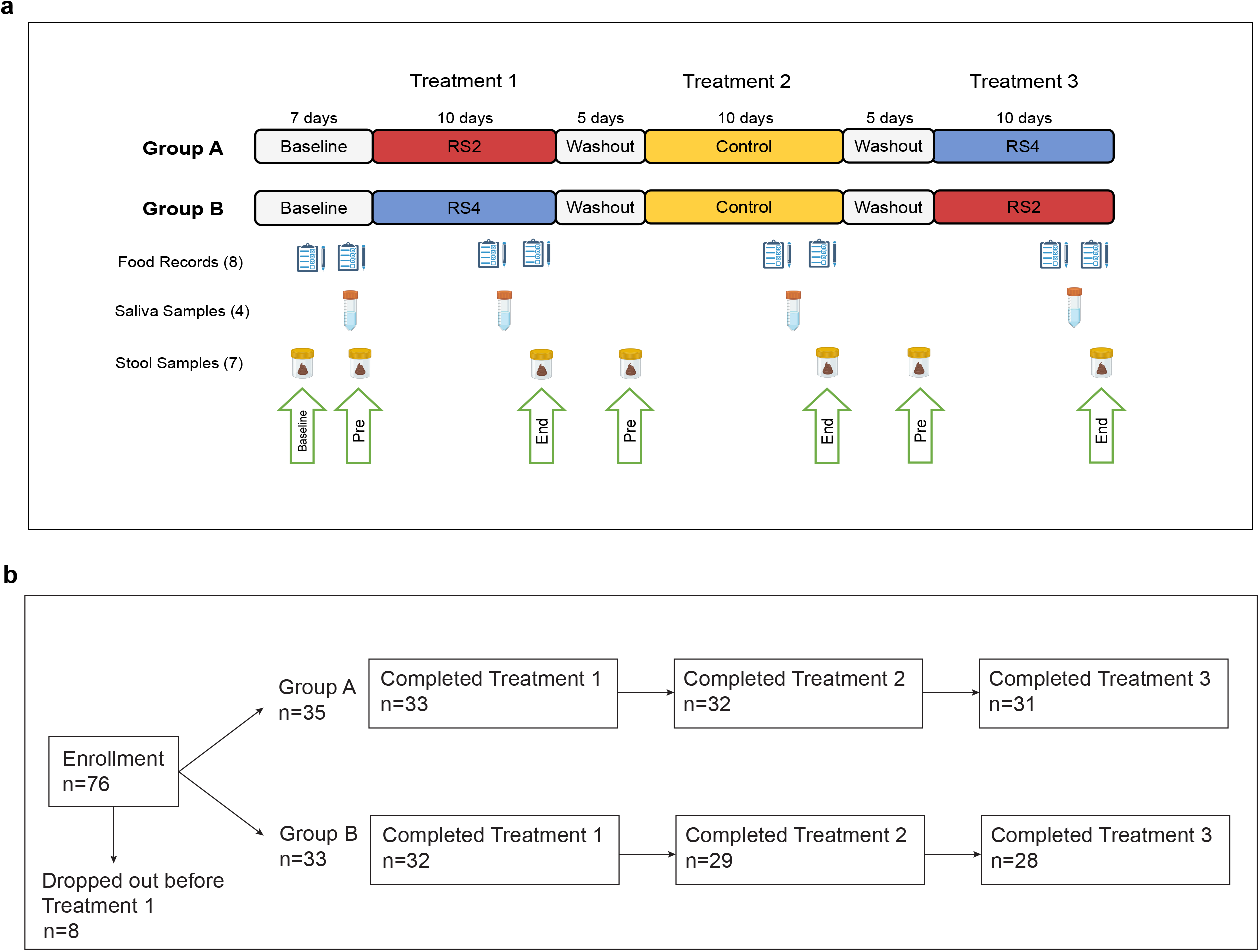
Study Design. (a) Participants were enrolled in one of two arms of a seven-week crossover dietary intervention study during which we supplemented their diets with crackers containing one of three starches: RS2 = resistant starch type 2, RS4 = resistant starch type 4, Control = digestible starch. (b) Flow diagram of participant enrollment and completion of treatments.

**Table 1.** Baseline characteristics for all participants assigned to a study arm. There were 35 participants in Group A and 33 in Group B, for a total of 68 who were assigned to a study arm. Data are presented as mean (standard deviation) or n (%). There were no significant differences between Group A and B at baseline for age, sex, body fat percentage for males, body fat percentage for females, and energy-adjusted dietary fiber intake at baseline. T-tests and Wilcoxon rank sum tests were used for numerical variables and chi-squared tests were used for binary variables.

|  | Group A<br>(n=35) | Group B<br>(n=33) | Total | *p* value |
| --- | --- | --- | --- | --- |
| Age in years | 28 (9) | 27 (11) | 27 (10) | 0.376 |
| Sex, n (%) |  |  |  | 0.542 |
| Female | 22 (62.9%) | 24 (72.7%) | 46 (67.6%) |  |
| Male | 13 (37.1%) | 9 (27.3%) | 22 (32.4%) |  |
| Body fat percentage |  |  |  |  |
| Female | 26.2 (7.9) | 24.9 (6.6) | 25.50 (7.2) | 0.531 |
| Male | 18.0 (5.1) | 20.40 (7.0) | 19.00 (5.9) | 0.351 |
| Energy-adjusted dietary fiber intake at baseline in grams | 12.7 (5.5) | 11.0 (4.3) | 11.9 (5.0) | 0.235 |

### RS, but not digestible starch, elicits an overall change in gut microbiome composition

For each treatment, we used MaAsLin2 models to assess overall microbial response, which was represented by ASVs that changed in relative abundance in the study cohort over the treatment period. We identified 34 ASVs that were differentially abundant between PreRS2 (before RS2) and EndRS2 (the end of RS2) treatment, 24 ASVs that were differentially abundant between PreRS4 and EndRS4, and no ASVs that were different between PreCtl (before control) and EndCtl (end of control) treatment (Supplementary Table 1). Of the 34 ASVs identified for RS2 and the 24 ASVs identified for RS4, we found 9 ASVs shared between the two treatments. Mean log_2_ fold changes of relative abundances of the differentially abundant ASVs identified by MaAsLin2 are shown in Figure 2A. Although we observed these population level shifts, we detected noticeable variation between individuals (Figure 2B and 2C). Measuring the degree of interindividual variability, we found that an ASV for *Ruminococcus bromii* (var=11.09) and an ASV for *Parabacteroides distasonis* (var=17.37) had the greatest variance in log_2_ fold change between the beginning and end of treatment with RS2 and RS4, respectively.

**Figure 2.**
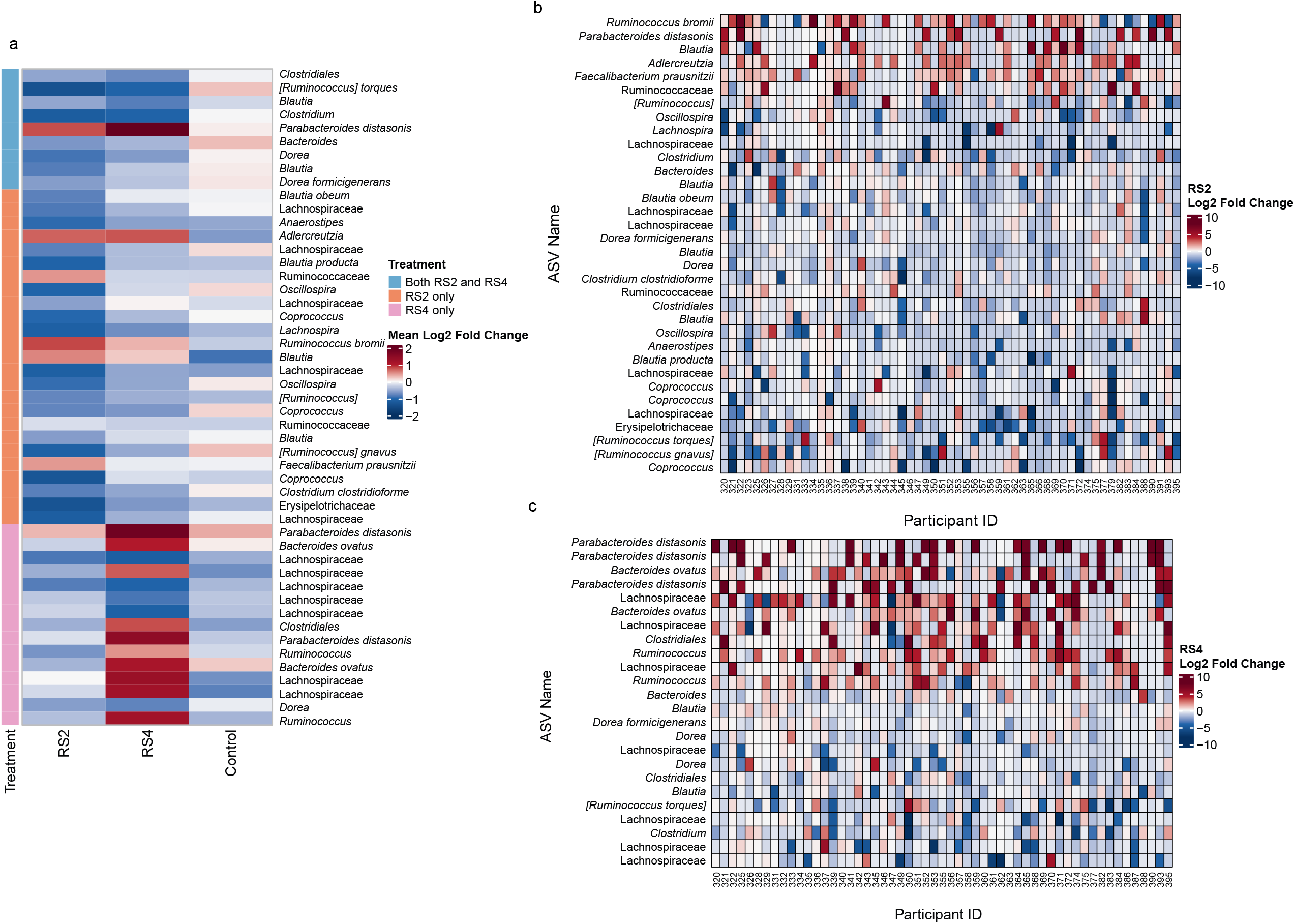
Change in gut microbiome composition after RS2 and RS4 treatments varies between individuals. (a) Heat map showing the change in mean relative abundance of ASVs across all participants by treatment (RS2, RS4, and control) for taxa that significantly differed in abundance from start to end of the RS2 and RS4 treatments in our MaAsLin2 models (q<0.05). No ASVs significantly changed in response to the control treatment. For each cell, colors indicate the log_2_ fold change in relative abundance of taxa between Pre treatment and End treatment time points 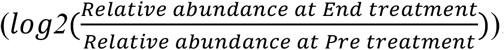. Color annotations on the left (ASVs significant) denote the treatment period during which the marked taxa were significantly differentially abundant between Pre and End of treatment. Repeated taxa indicate the same taxonomic assignment to different ASVs. (b,c) Log_2_ fold changes of the relative abundances of the ASVs for individual participants. ASVs included in these heat maps were identified to be significantly different between Pre and End of RS2 (b) or RS4 (c) by MaAsLin2.

### Dietary fiber intake and mean SAA are predictive of differential abundance of ASVs at the end of treatment periods

We used MaAsLin2 models to determine the ability of candidate predictors to predict differences in relative abundances between the Pre and End time points of the treatment periods. Our candidate predictors were: *AMY1* CN, *AMY1* CN Group (high: ≥ 9 copies versus low: ≤ 5 copies),^18^ mean SAA, dietary fiber intake during baseline, dietary fiber intake during treatment (excluding study cracker consumption), and treatment order (whether participants consumed RS2 or RS4 initially). The distribution of *AMY1* CN and mean SAA for individuals included in the analyses can be found in Figure 3A and Figure 3B. Of note, each candidate, except *AMY1* CN, had an interaction with time point for at least one ASV in at least one of the treatment periods at q<0.25 (Supplementary Table 2). We used the q<0.25 threshold here as this was a screening step. For the interaction terms with q<0.25, we next tested whether the corresponding candidate predictors could predict the relative abundance of ASVs at the End of each treatment period. High dietary fiber intake at baseline was predictive of lower relative abundance of *Ruminococcus torques* (q<0.03) and higher relative abundance of *Dialister* (q<0.03) at EndRS2, and high dietary fiber intake during the RS2 treatment period was predictive of higher relative abundance of *Coprococcus* at EndRS2 (q<0.04) (Table 2). High dietary fiber intake at baseline was predictive of lower relative abundance of *Oscillospira* at EndRS4 (q<0.002) (Table 2). Finally, at EndCtl, high mean SAA was predictive of higher abundance of *Sutterella* (q=0.04).

**Figure 3.**
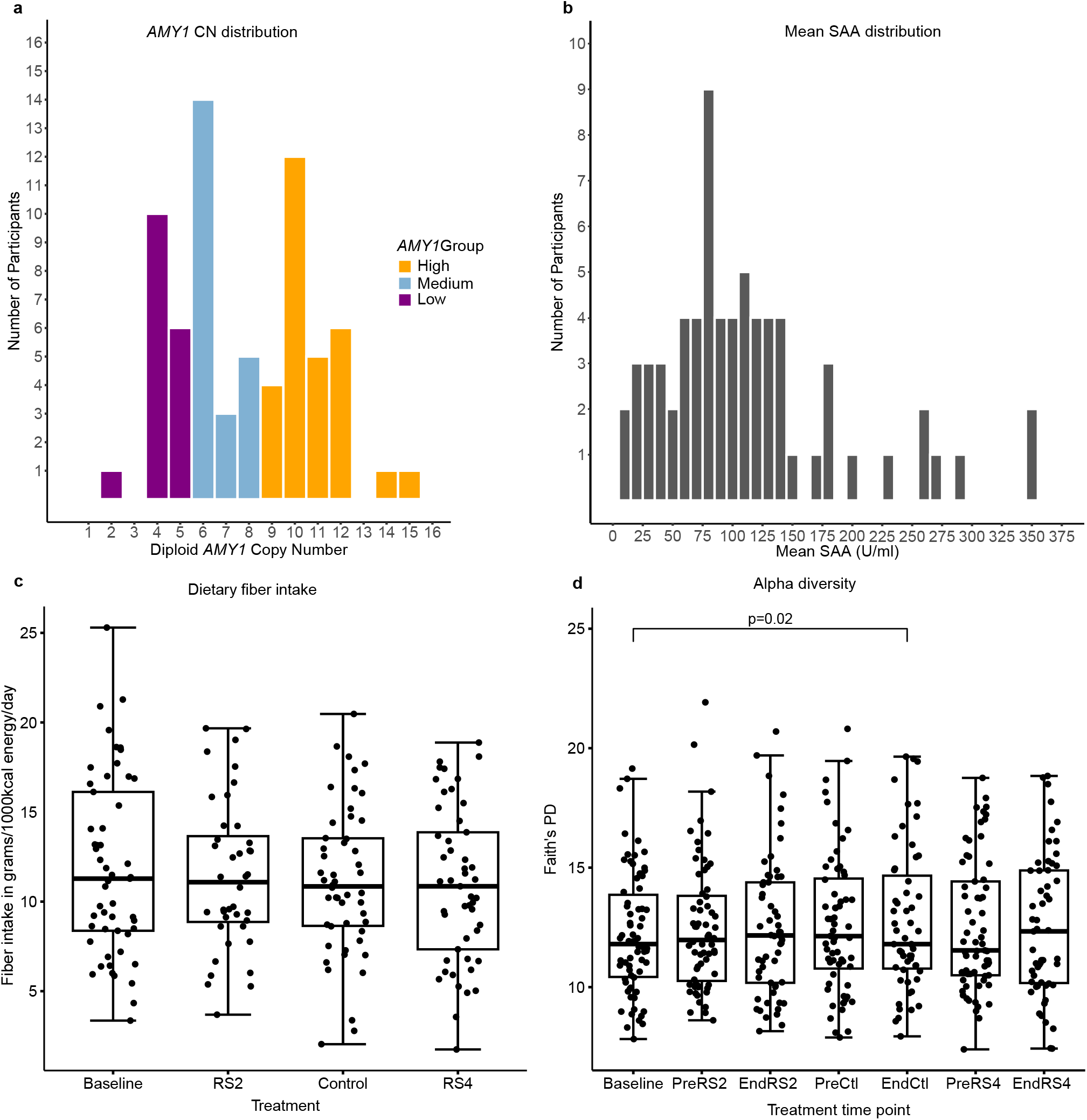
Characterization of candidate predictors in the study population. (a) *AMY1* CN and (b) mean SAA distribution in our study population for participants included in analyses. Low *AMY1* CN ≤ 5, High *AMY1* CN ≥ 9, and Medium *AMY1* CN ≥ 6 and ≤ 8. (c) There were no significant differences (p<0.05) in dietary fiber intake between the treatment periods. (d) We determined gut microbiome alpha diversity using Faith’s PD at each time point and tested for differences between time points using a linear mixed model. There were no significant differences in Faith’s PD between time points except a decrease observed at End Ctl compared to Baseline (ꞵ = -0.60, SE = 0.18, p = 0.02).

**Table 2.** Candidates that predict the relative abundance of ASVs at the End of treatment. Candidates that significantly predict the relative abundance of ASVs at the End of treatment at (q<0.05). The *β* column represents the change in relative abundance for the specified ASV for each unit of change in the specified candidate predictor variable. SE: standard error.

| Treatment | Candidate Predictor | ASV | $\beta$ | SE | *p* value | *q* value |
| --- | --- | --- | --- | --- | --- | --- |
| RS2 | Dietary fiber intake during Baseline | *[Ruminococcus] torques* | -1.37 | 0.33 | 0.0001 | 0.03 |
|  |  | *Dialister* | 0.49 | 0.12 | 0.0002 | 0.03 |
|  | Dietary fiber intake during RS2 | *Coprococcus* | 0.44 | 0.10 | 0.0001 | 0.04 |
| RS4 | Dietary fiber intake during Baseline | *Oscillospira* | -1.4 | 0.27 | $6.45 \times 10^{-6}$ | 0.002 |
| Ctl | Mean SAA | *Sutterella* | 1.3 | 0.31 | $9.99 \times 10^{-5}$ | 0.04 |

### Changes in fecal SCFA concentration are predicted by alpha diversity and treatment order

We found that, at the population level, both RS treatments decreased acetate, propionate, and total SCFA, while Ctl increased acetate, butyrate, propionate, and total SCFA (Figure 4A). However, we found interindividual variability in the observed SCFA concentration changes associated with each treatment period (Figure 4B). Using logistic regression models with SCFA change scores as the response variable, we found that treatment order predicted propionate response to RS2 (Group B: ꞵ=-2.63, p=0.0003) and RS4 (Group B: ꞵ=2.94, p=0.0004) and total SCFA response to RS4 (Group B: ꞵ=1.57, p=0.02) (Figure 4C). β coefficient estimates indicate the difference in the log-odds of a response (a change score of 1) between Group A and Group B. Alpha diversity, as measured by Faith’s PD, prior to RS2 treatment was a significant predictor of butyrate change score (ꞵ: -0.2504, p = 0.03) and acetate change score (ꞵ: -0.2518, p = 0.04) (Figure 5). In other words, for every unit increase in Faith’s PD at PreRS2, the odds of an increase in butyrate and acetate were each 22% lower.

**Figure 4.**
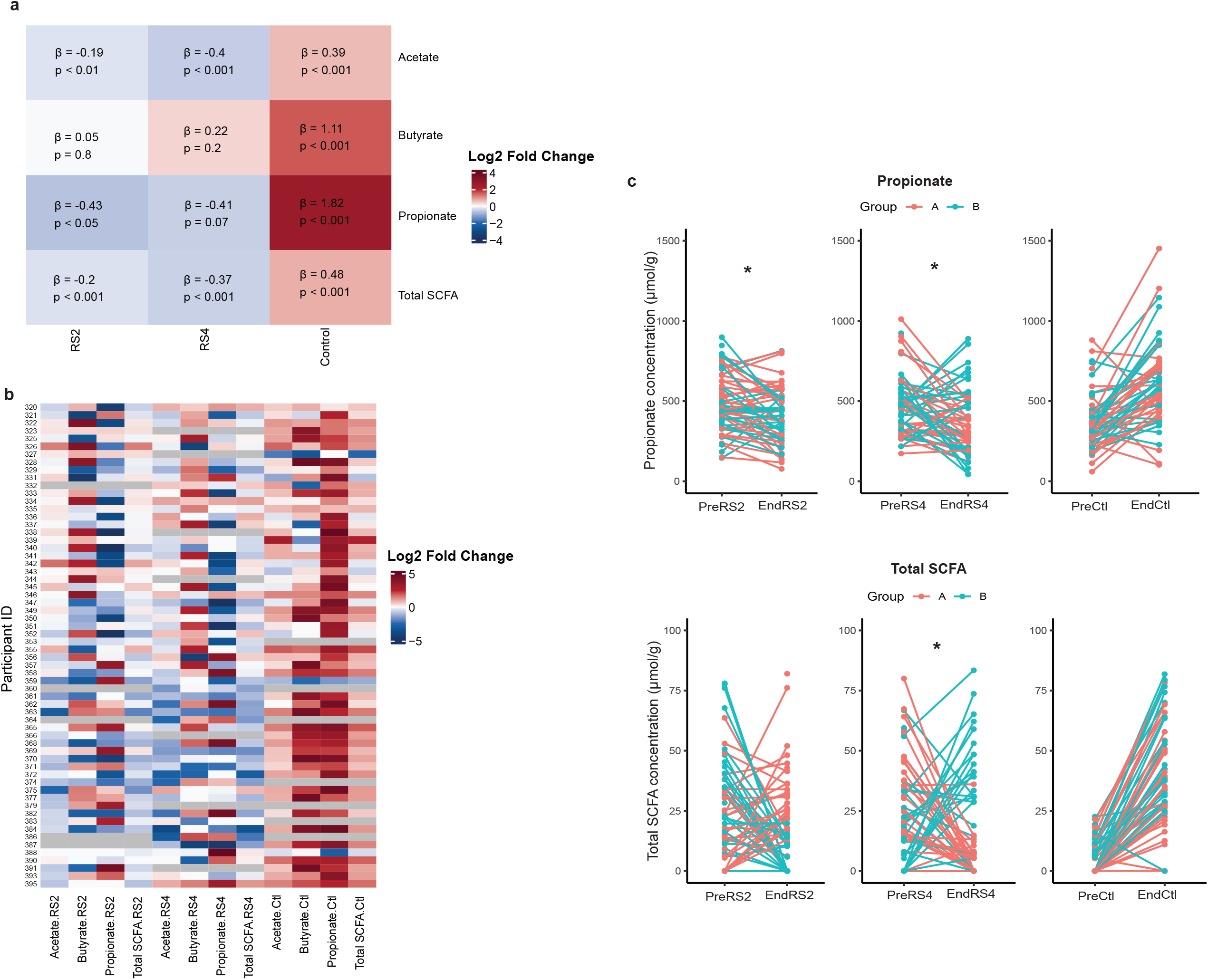
Conserved and variable fecal SCFA response to RS2, RS4 and control treatments. (a) This heat map depicts the results of linear mixed models with the time point (Pre vs End) of each treatment as a fixed effect, participant as a random effect, and log SCFA concentration as the response variable. Coefficient estimates (*β*) indicate whether the specified SCFA increased or decreased at End compared to Pre for each treatment. (b) We observe interindividual variability in log_2_ fold changes of fecal SCFA concentrations between Pre and End of each treatment. (c) Spaghetti plots shown here depict change in propionate and total SCFA concentrations between Pre and End of each treatment period for individual participants. Panels are marked with an asterisk (*) where treatment order (see Figure 1A) was a significant predictor of change score as determined by logistic regression analysis.

**Figure 5.**
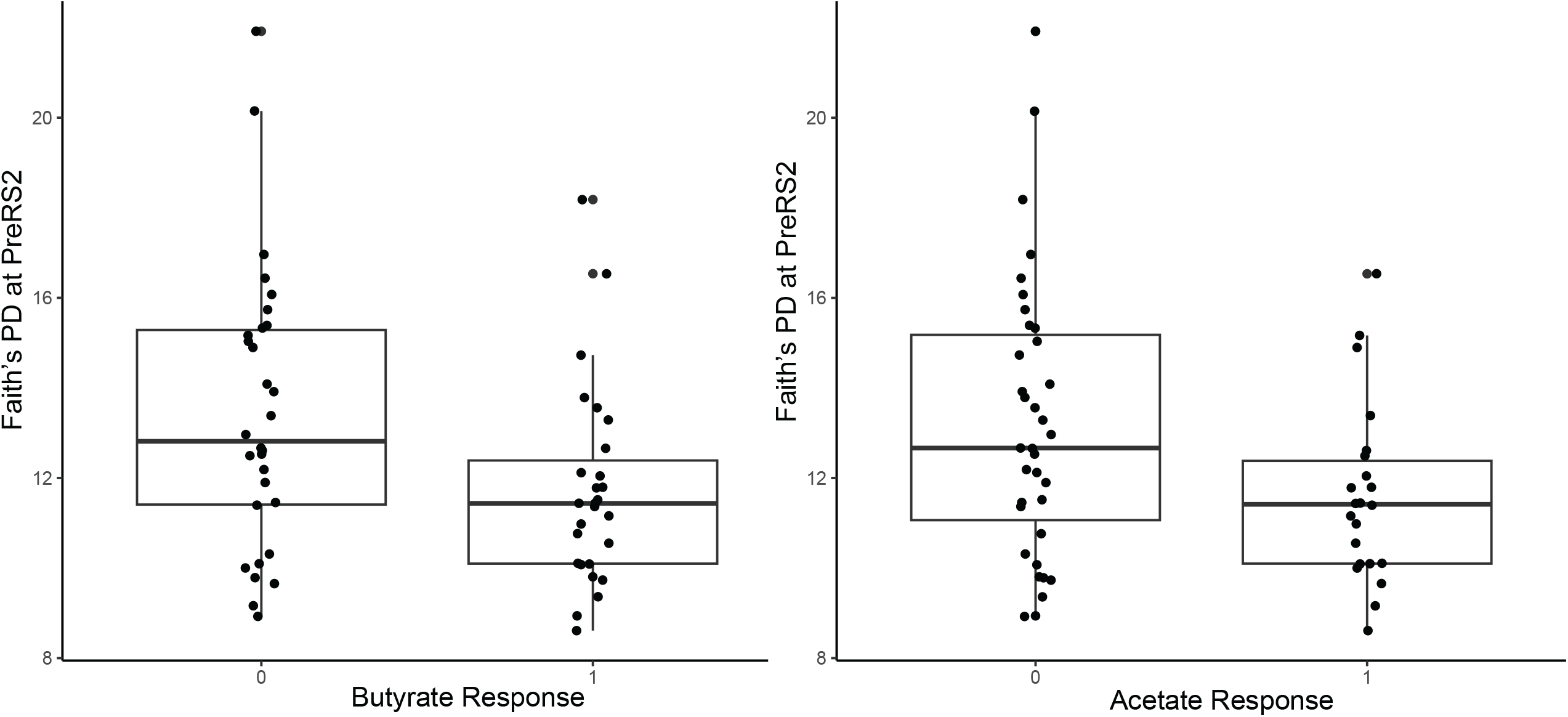
Gut microbiota diversity prior to RS2 treatment predicts change in butyrate and acetate in response to RS2 treatment. Faith’s PD prior to the consumption of RS2 is a significant predictor of butyrate change score (ꞵ=-0.2504, p=0.03) and acetate change score (ꞵ=-0.25, p=0.04) as determined by logistic regression analysis.

### Relative abundances of ASVs at PreRS2 predict propionate response to RS2

We performed LASSO regression to select the most important variables, including ASVs at Pre, that predict butyrate, propionate, or acetate response to each treatment. We included ASVs at Pre as variables in our models to determine which microbes present in individuals prior to each treatment could be predictive of SCFA response. Similar to our logistic regression models, when applying LASSO, the predictor variables were most successful in predicting propionate response (Table 3). We found that 14 variables, treatment order and the relative abundances of 13 ASVs at PreRS2, were predictive of propionate response, defined as the log fold change of propionate between PreRS2 and EndRS2, with R^2^ = 0.64. Additionally, two variables, treatment order and the relative abundance of *Clostridium symbiosum* at PreRS4, were predictive of propionate response to RS4 (R^2^ = 0.31). No ASVs were predictive of change in acetate. Full results are presented in Table 3.

**Table 3.** ASVs at Pre RS and treatment order are predictive of propionate response to RS. This table shows the contributions of independent variables included in LASSO models to predict the log fold change of propionate and butyrate concentrations, as represented by their *β* coefficient values, as well as the percentage of the dependent variable explained by the independent variables (**R**^**2**^). The relative abundances of the ASVs at Pre of the specified treatment period were used in the models. Repeated taxa within the same treatment indicate the same taxonomic assignment for different ASVs. Empty cells indicate the specified variable did not contribute to the model.

| Treatment | Predictor | Response:<br>Propionate | Response:<br>Butyrate |
| --- | --- | --- | --- |
| RS2 | Treatment Order | -1.58 |  |
|  | Lachnospiraceae | 69.35 |  |
|  | *Oscillospira* | 2428.80 |  |
|  | *Anaerostipes* | -19.98 |  |
|  | *Bacteroides plebeius* | -0.39 |  |
|  | *Clostridium symbiosum* | 272.62 |  |
|  | *Coprococcus* | 3.54 |  |
|  | Oscillospira | 381.87 |  |
|  | Lachnospiraceae | 31.31 |  |
|  | *Alistipes massiliensis* | -23.35 |  |
|  | Lachnospiraceae | 75.50 |  |
|  | *Lactococcus* | 33.15 |  |
|  | *Ruminococcus* | -109.28 |  |
|  | *Ruminococcus* | -7.16 |  |
|  | *Coprococcus* |  | 4.30 x 10<sup>-15</sup> |
|  |  | **R<sup>2</sup>=0.64** | **R<sup>2</sup>=0** |
| RS4 | Treatment Order | 1.15 |  |
|  | *Clostridium symbiosum* | 31.69 |  |
|  |  | **R<sup>2</sup>=0.31** | **R<sup>2</sup>=0** |
| Treatment | Predictor | Response:<br>Propionate | Response:<br>Butyrate |
| Control | *Bacteroides caccae* |  | -6.74 |
|  | AMY1 Group |  | 0.07 |
|  |  | **R<sup>2</sup>=0** | **R<sup>2</sup>=0.06** |

## Discussion

In this study, we identified predictors of gut microbiota response to two types of resistant starch, RS2 and RS4, and a digestible corn starch. We found that dietary fiber intake and mean SAA were the most significant predictors of changes in relative abundances of ASVs following RS and control treatments (Table 2). We also observed that treatment order and pretreatment gut microbiome alpha diversity significantly predicted fecal SCFA responses (Figure 4C and Figure 5).

Most of the significantly changed ASVs by the end of RS treatments were decreased in relative abundance, particularly for RS2 (Figure 2). Decreases in relative abundance after a dietary intervention are not necessarily surprising, because only the microbes that use the given substrate might be preferentially enriched over those that do not. In fact, the ASVs that were most significantly increased at EndRS2 corresponded to *R. bromii* and *Blautia*. These changes could be indicative of a shift towards increased capacity for SCFA production because *R. bromii* is a primary resistant starch degrader that breaks down starch into intermediate metabolites used by butyrate-producing microbes, and the *Blautia* genus plays a role in the production of acetate and propionate.^26–28^ Nevertheless, we also observed interindividual variability in relative abundances after both RS2 and RS4 consumption, in accordance with previous studies.^6,10^ Although two ASVs were increased at the population level, *R. bromii* during RS2 (q=0.044) and *P. distasonis*, which has beneficial effects,^29^ during RS4 (q<0.001), these ASVs also had the greatest variance in relative abundance between individuals. Consequently, determining factors that predict individual response in terms of these bacteria may be relevant to whether an individual can reap health benefits from consuming RS2 or RS4.

We found that prior dietary fiber intake could impact an individual’s response to RS supplementation, and the outcome differed depending upon the type of RS consumed (Table 2). This result suggests that dietary history should be considered when manipulating gut microbiome composition to promote beneficial species or decrease pathogenic species. This result is supported by our finding that treatment order also predicts SCFA response to RS supplementation (Figure 4C).

We previously found that people with a high *AMY1* CN have a greater abundance of gut microbes that ferment dietary fibers, such as RS, than people with a low *AMY1* CN.^18^ Thus, we posited that individuals with high *AMY1* CN or SAA would have greater enrichment of microbes that degrade RS and greater production of SCFAs resulting from RS supplementation compared to individuals with low *AMY1* CN or SAA.^18^ Contrary to this, the only treatment after which an *AMY1*-related metric was significantly predictive of relative abundance was the control starch, which is digestible by host enzymes including salivary amylase. We found that mean SAA was positively correlated (Pearson’s r=0.46, p<0.001) with relative abundance of *Sutterella* at EndCtl (Table 2). *Sutterella*, an asaccharolytic microbe incapable of using polysaccharides as an energy source, is increased in individuals with metabolic syndrome on a low fiber diet compared to a dietary fiber- and RS-enriched diet.^30^ Our finding could be explained by competition between host and gut microbes for digestible substrate. With decreased availability of digestible starch because of host amylase degradation, the intestinal environment favors *Sutterella*. Interestingly, *Sutterella* and SCFAs have been observed in a positive feedback relationship. Peterson and colleagues found that, *in vitro*, increasing SCFAs can modulate an increase in *Sutterella*, speculating that the bacterium might have evolved to use SCFAs for energy due to its inability to use polysaccharides.^31^ Although *AMY1* CN was not a strong predictor of response to RS at the ASV level in this study, analysis of shotgun metagenomics data is warranted to determine the ability of *AMY1* CN to predict changes in carbohydrate-active enzymes in response to RS.^32^

At the population level, RS2 and RS4 treatments decreased SCFA concentrations by the End of each treatment compared to Pre (Figure 4A). These findings were surprising given our hypothesis that RS2 and RS4 would overall be associated with the enrichment of SCFA-producing microbes and, in turn, increased SCFA concentrations. However, the interindividual variability in response to RS consumption that we observed would provide a plausible explanation for this result (Figure 4B). Further supporting the role of interindividual variability in our observations, studies on the impact of RS supplementation on host physiological outcomes, such as improvements in fasting blood glucose and insulin concentrations, have been conflicted.^33–35^ Because microbially generated SCFAs are involved in signaling pathways that impact blood glucose concentrations, these conflicts regarding the effectiveness of RS at improving glucose homeostasis may be due, at least in part, to interindividual variability in gut microbiota response to the treatment.^1,36,37^ Future work would be required to determine the effectiveness of our predictors at predicting host physiological outcomes.

The most striking result was that treatment order (the order in which participants received RS2 and RS4) was a significant predictor of SCFA increase at End compared to Pre. Specifically, participants who received RS2 first (Group A) were more likely to have a propionate response to RS2 compared to those who received RS4 in the first treatment period (Group B). Conversely, participants in Group B were more likely to have a propionate response and a total SCFA response to RS4 compared to Group A, who received RS2 first. Taken together, these results indicate a greater likelihood of a response to the RS received as the first of the three treatments during the study. The lack of a response during the third treatment, during which Group B switched to RS2 and Group A switched to RS4, could be suggestive of a priming effect from the first or second treatment on the microbial composition that carried over to the third treatment period. We speculate that the first treatment caused changes in the community’s functional capacity, which enhanced the efficiency of RS2 degradation in Group A and RS4 degradation in Group B. Finally, we found that lower alpha diversity prior to RS2 consumption was associated with higher odds of acetate and butyrate responses to RS2. The relationship between lower diversity and acetate and butyrate responses to RS2 may be due to the enrichment of microbes that ferment RS2 or produce SCFAs with the expense of a decrease in other microbes.

Our study does have some limitations. The treatments could have caused the participants to adjust their dietary intake, e.g., the study crackers could have replaced another component of their usual diet or may have increased their overall caloric intake, which could have modified the effects attributed to the treatments. This is an inherent challenge of any dietary intervention study in which participants can consume their usual diets ad libitum. Nevertheless, the advantages of our approach were that participants were not asked to abandon their preferred habitual intake and the RS was provided in a purified form as opposed to being added in the form of RS-rich whole foods, which assured uniformity in supplementation of the RS types. Additionally, as in all studies that use measurement of SCFA concentrations in stool to quantify microbial community output, we were limited to capturing the 5-10% of SCFAs that are excreted in stool. This measure is typically used as a proxy for the amount of SCFAs produced by colonic microbes in microbiome studies because more accurate procedures, such as sampling hepatic portal blood, can be impractical for human studies.

Individualized gut microbial responses have the potential to influence the metabolic effects of the consumption of dietary fibers such as RS. In this study, we identified several factors that predict the variable response of gut microbiota to RS2 and RS4. Our findings support the premise that a personalized approach via characterizing features, such as host dietary fiber consumption and gut microbiota composition, prior to a nutritional intervention with RS may increase the likelihood of favorable health outcomes like increased beneficial SCFA production in the gut.

## Methods

### Study participants

This study was conducted at Cornell University in Ithaca, New York. All human-related procedures and sample and data collection were approved by the Cornell University Institutional Review Board for Human Participant Research (Protocol Number: 1902008575) prior to recruitment and enrollment of participants. Study participants included healthy males and healthy, non-pregnant or lactating females 18-59 years old. Baseline participant characteristics can be found in Table 1. Exclusion criteria included a history of gastrointestinal diseases or surgeries; type 1 or type 2 diabetes, prediabetes or impaired glucose tolerance, self-reported untreated thyroid condition; use of antibiotics 6 months prior to the start of the study; and chronic alcohol intake (>5 drinks/day). This trial was registered at clinicaltrials.gov as NCT05743790 on February 24, 2023.

### Study design

We enrolled eligible participants into a seven-week open-label crossover dietary intervention study (Figure 1A). Each participant was assigned a study identification (ID) number based on the order in which they were enrolled in the study. All participants with an odd ID number were allocated to Group A, and all participants with an even ID number were allocated to Group B. A single participant with an odd ID number was allocated to Group B to balance the groups.

Participants were instructed to consume crackers containing RS2, RS4, or a digestible control starch (Ctl), in their assigned treatment order, each for 10 days (Figure 1), in addition to their normal dietary intake. The groups received treatments in the following order. Group A: Treatment 1 = RS2, Treatment 2 = Ctl, Treatment 3 = RS4; Group B: Treatment 1 = RS4, Treatment 2 = Ctl, Treatment 3 = RS2. The three treatment periods were separated by 5-day washout periods when no study crackers were consumed. Participants collected fecal samples prior to (“PreRS2,” “PreCtl,” or “PreRS4”) and at the end of (“EndRS2,” “EndCtl,” or “EndRS4”) the treatment periods (Figure 1: see “Pre”and “End” time points).

### Dietary supplementation

Participants were provided with crackers that contained RS2 (HI-MAIZE® 260 starch, Ingredion), RS4 (VERSAFIBE™ 1490 starch, Ingredion), or a digestible control starch (AMIOCA™ TF starch, Ingredion). Study cracker formulations are listed in Supplementary Table 3. During treatments 1 and 3, we aimed to provide 30 g of each type of RS per day. Following production, the crackers were analyzed for RS and dietary fiber content by Medallion Laboratories (Minneapolis, MN) using AOAC 2002.02 and AOAC 991.43 methods, respectively (Supplementary Table 4). After baking, the amount of RS in RS2 crackers was reduced from 30 g to 21.27 g per serving, possibly due to starch gelatinization. During each 10-day treatment period, we followed a dose escalation of study crackers where participants gradually increased the dosage over four days from the beginning of treatment to a final dose of 21.27 g per day for the last 7 days (Day 1: 25%, Day 2: 50%, Day 3: 75%, Day 4 - Day 10: 100%). Participants were asked to maintain their habitual dietary intake and physical activity across interventions and to avoid taking prebiotic- or probiotic-added foods, drinks, or supplements throughout the study.

Study crackers were provided in individual bags containing a preportioned daily supply by weight. At the end of each treatment period, participants were asked to complete a short questionnaire to indicate the percentage of crackers consumed per day as an indicator of adherence to the protocol. Data from participants who reported a low adherence to cracker consumption were excluded from analysis. Low adherence was defined as having consumed <75% of study crackers for five or more days during the treatment or <50% on the day of or day before collecting their End time point fecal sample.

### Anthropometric measurements

At baseline, we measured the participants’ total body fat percentage with a Tanita SC-240 Total Body Composition Analyzer using Bioelectrical Impedance Analysis (BIA) as per the manufacturer’s instructions.

### Dietary intake data

During the baseline and treatment periods, we instructed participants to complete two nonconsecutive 1-day food records of one weekday and one weekend day, excluding study cracker consumption, using the Automated Self-Administered 24-hour (ASA24) Dietary Assessment Tool 2020 developed by the National Cancer Institute, Bethesda, MD (https://epi.grants.cancer.gov/asa24).^38^ We adjusted dietary fiber intake by energy (g/1,000 kcal/day) to account for differences in participants’ overall energy intake. Data were averaged from the two food records for each participant within the baseline week and each of the three treatments. Data were excluded if only one of the two records was completed.

### Saliva collection

We obtained 5 ml of saliva at baseline and during each treatment for a total of four saliva samples throughout the study for each participant (Figure 1). Participants were instructed to refrain from brushing their teeth for a minimum of six hours and to avoid consuming any food or beverages including water for a minimum of 30 minutes prior to sample collection. They were then instructed to accumulate saliva in their mouth and express it into a 50 ml sterile conical tube. Saliva samples were stored on ice immediately after collection, aliquoted within three hours, and stored at -80°C.

### AMY1 copy number determination by qPCR and ddPCR

Genomic DNA was extracted from saliva samples using the QIAamp 96 DNA Blood Kit, QIAamp Blood Mini Kit, and the QIAamp Investigator Kit (Qiagen, cat # 51161, 51104, 56504). We performed qPCR using primers, previously described in Poole et al.,^18^ to amplify *AMY1* paralogs and our reference gene, *EIF2B2* (CN=2). For each gene, each qPCR reaction consisted of 1 μl genomic DNA normalized to 5 ng/μl, 0.5 μl of each forward and reverse primer at 10 μM, 3 μl of PCR grade H2O, and 5 μl iTaq™ Universal SYBR® Green Supermix (BioRad, cat # 1725122), for a total volume of 10 μl per reaction. The qPCR conditions were as follows: initial denaturation at 95°C for 5 minutes and 40 cycles of 95°C for 10 seconds and 60°C for 30 seconds on a Roche LightCycler 480 Real-Time PCR Instrument. We made a standard curve using genomic DNA NA12286 (Coriell Institute; *AMY1* CN = 2). The following genomic DNAs were used as positive controls on all qPCR plates, as their *AMY1* copy numbers have previously been estimated: NA18972, NA12873, NA10472, NA12890, NA10852, NA12043, NA11992, NA12414, NA12340, NA06994, NA12342, NA12286, NA18522, and NA19138 (Coriell Institute). All reactions including standards and blanks were performed in quadruplicate and results were averaged for technical replicates with a coefficient of variation <0.05. At least two qPCR runs were performed for all participants and we calculated the median value of all qPCR results to determine the final qPCR *AMY1* CN value. For digital PCR, genomic DNA was digested with the restriction enzyme *Hae*III (New England Biolabs, cat # R0108S) and diluted to a final concentration of approximately 15 ng/µl. Digital PCR was performed using Life Technologies Taqman Copy Number Assay Id Hs07226361_cn for the *AMY1* locus and TaqMan Copy Number Reference Assay Hs06006763_cn for the AP3B1 gene to normalize for total DNA. The reactions were run on a QX100 Droplet Digital PCR System in duplicate. We averaged the median of all qPCR results and the two digital PCR results to determine the final *AMY1* CN used in our analyses.

### Salivary amylase activity assay

We measured SAA for each saliva sample in triplicate, from up to four saliva samples donated from each participant throughout the duration of the study, using the Salimetrics Salivary Alpha-Amylase Enzymatic Kit (SALIMETRICS, cat # 1-1902). The manufacturer’s protocol was followed except for the use of 300 µl amylase substrate per reaction instead of 320 µl. We averaged SAA determined for each participant to obtain the mean SAA used in our analyses.

### Fecal sample collection

Participants collected fecal samples at seven time points throughout the study (Figure 1). Samples were collected from a single bowel movement and stored at -80°C within 24 hours of collection. We lyophilized an aliquot from each sample for 16S rRNA gene sequencing. Samples from study participants who reported collection of fecal samples ≥ two days after completion of the treatment were excluded from analysis.

### 16S rRNA gene sequencing

DNA extraction was performed on 0.030–0.045 g lyophilized fecal samples using the DNeasy PowerSoil 96 HTP Kit (Qiagen, cat # 12888-100), following the manufacturer’s instructions with the specifications: samples were loaded into PowerBead Plates and stored at -20 °C in Bead Solution until extraction, and instead of vortexing to mechanically lyse samples, samples were placed in a BioSpec 1001 Mini-Beadbeater-96 for three minutes. We amplified the V4 region of the 16S rRNA gene using the universal primers 515F and barcoded 806R^39^ and approximately 100 ng of genomic DNA from each sample in duplicate PCR reactions using 25 µl Classic++™ Hot Start Taq DNA Polymerase Master Mix (Tonbo Biosciences, cat # TB-31-5011-1000R), 22 µl PCR water, 0.5 µl of each 10 nM primer, and 2 µl DNA. We used the PCR program previously described but with 25 cycles of amplification.^39^ We purified amplicons using Mag-Bind TotalPure (Omega Bio-tek, cat # M1378-01) using a 1.8X bead ratio, pooled 100 ng of amplicons from each sample, and performed 2×250bp sequencing on an Illumina MiSeq instrument. We performed microbiome bioinformatics with QIIME 2.^40^ We demultiplexed and quality filtered raw sequence data via q2-demux, denoised with DADA2^41^ via q2-dada2, aligned ASVs with mafft^42^ via q2-alignment, and constructed a phylogenetic tree using fasttree^43^ via q2-phylogeny. We used q2-diversity to calculate alpha-diversity (Faith’s Phylogenetic Diversity, Faith’s PD)^44^ after samples were subsampled without replacement to 23081 sequences per sample based on the sample with the lowest sequence count. We used the q2-feature-classifier^45^ classify-sklearn naïve Bayes classifier against the Greengenes 13_8 99% OTUs reference sequences^46^ to assign taxonomy to ASVs.

### Short-chain fatty acid measurements

SCFAs including acetate, propionate, isobutyrate, butyrate, isovalerate, valeric acid, isocaproic acid, caproic acid, and heptanoic acid were quantified using ultra-performance liquid chromatography (Acquity UPLC system, Waters Corporation, Milford, MA) at the PennCHOP Microbiome Program Microbial Culture & Metabolomics Core. Total SCFA concentration was calculated as the sum of all nine SCFAs quantified.

### Statistical analysis

We performed all statistical analyses using RStudio version 4.2.1.^47^ We considered p-values < 0.05 to be statistically significant. When adjusting p-values we used the Benjamini–Hochberg false discovery rate correction and considered a q-value (adjusted p-value) of <0.05 to be statistically significant. To calculate fold changes of ASVs, we added a pseudocount of 1 to all ASV counts prior to calculating relative abundances. We used R package ComplexHeatmap^48^ and circlize^49^ to generate all heat maps.

To assess the response of ASVs to each treatment, we used the R package MaAsLin2 (Microbiome Multivariable Association with Linear Models 2).^50^ We normalized ASVs using total sum scaling then fit log-transformed linear mixed models after 10% prevalence filtering. We fit three types of models. First, we fit main effects models using time point (PreRS2 vs EndRS2, PreRS4 vs EndRS4, or PreCtl vs EndCtl) as a fixed effect and participant as a random effect in each model to identify ASVs that, on average, changed in relative abundance from Pre to End of each treatment period. Then we added interaction terms to each of these separate models (*AMY1* CN x time point, *AMY1* Group x time point, mean SAA x time point, baseline dietary fiber intake x time point, treatment dietary fiber intake x time point, and treatment order x time point), to evaluate the ability of the candidate predictors to affect changes in relative abundances. We also fit main effects models at End time points only, with each candidate predictor as an independent variable in a separate model, to detect whether they were predictive of relative abundance of ASVs at the end of each treatment.

For SCFA analysis we fit linear mixed models with log SCFA concentration as the response variable, time point (Pre vs End) as a fixed effect and participant as a random effect. We also fit logistic regression models with SCFA change score as the response variable (1= increase in concentration and 0 = decrease or no change in concentration from Pre to End of each treatment period), and treatment order and Faith’s PD as fixed effects in separate models. Since the limit of detection of the quantification method was 5.0 µmol/g of stool, a count of 2.5 µmol/g was added to all raw SCFA measurements for analyses where fold change was calculated.

We used the R package glmnet^51^ to perform 10-fold cross-validated Least Absolute Shrinkage and Selection Operator (LASSO) linear regressions to model the log fold change of acetate, propionate, and butyrate concentrations between Pre and End of each treatment period. For variable selection, we included the predictors: 10% prevalence-filtered ASVs at Pre, body fat percentage, sex, treatment order, *AMY1* CN, *AMY1* Group, mean SAA, and self-reported physical activity (minutes/week of vigorous, moderate, and low intensity activity) at baseline.

## Data Availability

The raw fastq sequences generated from the 16S rRNA amplicon sequencing will be available in the National Center for Biotechnology Information Sequence Read Archive via the project number PRJNA944997 upon publication of the manuscript in a peer-reviewed journal.

http://www.ncbi.nlm.nih.gov/bioproject/944997

## Acknowledgements

The authors’ responsibilities were as follows—ACP and SD: designed the research; SD, DKS, and JR: conducted the research; SD, DKS, AL, and LMJ: analyzed the data; DKS, SD, and ACP: wrote the paper; ACP had primary responsibility for final content; and all authors: reviewed and approved the manuscript prior to submission. The authors report there are no competing interests to declare. Research reported in this publication was supported in part by a President’s Council of Cornell Women Award (ACP) and the National Institutes of Health under award T32-DK007158 (DKS). The content is solely the responsibility of the authors and does not necessarily represent the official views of the President’s Council of Cornell Women or the National Institute of Diabetes and Digestive and Kidney Diseases or the National Institutes of Health. The resistant starches (HI-MAIZE® 260 starch, VERSAFIBE™ 1490 starch) and control starch (AMIOCA™ TF starch) were provided free of cost from Ingredion Inc. (Bridgewater, NJ). There was no industrial involvement in the design of this study or the interpretation of the data. This study used DNA from human lymphoblastoid cell lines from the NHGRI Sample Repository for Human Genetic Research and NIGMS Human Genetic Cell Repository of the Coriell Institute for Medical Research. We thank the Biotechnology Resource Center Genomics Facility at the Cornell Institute of Biotechnology for performing sequencing. Figure 1A was created in part using icons from BioRender.com. We also thank Jiayuan Liu, Jaden Ombres, Jacqueline Anaeto, and Belle Lin for their assistance. Finally, we thank our study participants without whom this research would not have been possible.

## Data availability

The raw fastq sequences generated from the 16S rRNA amplicon sequencing will be available in the National Center for Biotechnology Information Sequence Read Archive via the project number PRJNA944997 upon publication in a peer-reviewed journal.

