## Supplemental tables for "Gut microbial features and dietary fiber intake predict gut microbiota response to resistant starch supplementation"

**Supplementary Table 1.** Significantly different ASVs between Pre and End of each treatment period as determined by MaAsLin2

| **Treatment** | **ASV** | **Coefficient** | **SE** | ***p* value** | **q value** |
| --- | --- | --- | --- | --- | --- |
| RS2 | *Blautia* | -0.88 | 0.10 | <0.001 | <0.001 |
|  | *Coprococcus* | -0.68 | 0.11 | <0.001 | <0.001 |
|  | *Adlercreutzia* | 0.66 | 0.13 | <0.001 | <0.001 |
|  | *Dorea* | -0.79 | 0.15 | <0.001 | <0.001 |
|  | Erysipelotrichaceae | -1.3 | 0.24 | <0.001 | <0.001 |
|  | *Blautia obeum* | -0.72 | 0.14 | <0.001 | <0.001 |
|  | *Dorea formicigenerans* | -0.56 | 0.12 | <0.001 | <0.001 |
|  | Lachnospiraceae | -0.73 | 0.15 | <0.001 | <0.001 |
|  | *[Ruminococcus] torques* | -1.1 | 0.23 | <0.001 | <0.001 |
|  | *Clostridium clostridioforme* | -0.72 | 0.16 | <0.001 | 0.001 |
|  | *Coprococcus* | -0.54 | 0.12 | <0.001 | 0.002 |
|  | *Clostridium* | -1.0 | 0.24 | <0.001 | 0.002 |
|  | *Blautia* | -0.55 | 0.14 | <0.001 | 0.003 |
|  | *Parabacteroides distasonis* | 0.60 | 0.15 | <0.001 | 0.004 |
|  | *Oscillospira* | -0.65 | 0.17 | <0.001 | 0.008 |
|  | Clostridiales | -0.56 | 0.15 | <0.001 | 0.01 |
|  | *Oscillospira* | -0.73 | 0.21 | <0.001 | 0.01 |
|  | Ruminococcaceae | 0.71 | 0.20 | <0.001 | 0.02 |
|  | Lachnospiraceae | -0.88 | 0.26 | 0.001 | 0.02 |
|  | *[Ruminococcus]* | -0.71 | 0.21 | 0.001 | 0.02 |
|  | *Blautia* | -0.42 | 0.13 | 0.002 | 0.02 |
|  | *Coprococcus* | -0.52 | 0.16 | 0.0021 | 0.03 |
|  | *Anaerostipes* | -0.37 | 0.12 | 0.003 | 0.04 |
|  | *Lachnospira* | -0.65 | 0.21 | 0.003 | 0.04 |
|  | *Blautia* | 0.79 | 0.26 | 0.003 | 0.04 |
|  | Lachnospiraceae | -0.32 | 0.10 | 0.003 | 0.04 |
|  | *[Ruminococcus] gnavus* | -0.83 | 0.27 | 0.003 | 0.04 |
|  | Lachnospiraceae | -0.66 | 0.22 | 0.004 | 0.04 |
|  | *Blautia producta* | -0.52 | 0.18 | 0.004 | 0.04 |
|  | Lachnospiraceae | -0.30 | 0.10 | 0.004 | 0.04 |
|  | *Ruminococcus bromii* | 1.1 | 0.34 | 0.004 | 0.04 |
|  | Ruminococcaceae | 0.38 | 0.13 | 0.004 | 0.04 |
|  | *Faecalibacteriumprausnitzii* | 0.46 | 0.15 | 0.004 | 0.04 |
|  | *Bacteroides* | -0.50 | 0.17 | 0.005 | 0.04 |
| RS4 | *Parabacteroides distasonis* | 0.95 | 0.18 | <0.001 | 0.0001 |
|  | *Ruminococcus* | 1.28 | 0.23 | <0.001 | 0.0001 |
|  | *Blautia* | -0.66 | 0.12 | <0.001 | 0.0001 |
|  | Lachnospiraceae | 1.0 | 0.23 | <0.001 | 0.004 |
|  | *Parabacteroides distasonis* | 0.77 | 0.19 | <0.001 | 0.005 |
|  | *Parabacteroides distasonis* | 0.85 | 0.21 | <0.001 | 0.005 |
|  | Lachnospiraceae | -0.66 | 0.16 | <0.001 | 0.006 |
|  | Lachnospiraceae | 1.5 | 0.37 | <0.001 | 0.007 |
|  | Clostridiales | -0.65 | 0.16 | <0.001 | 0.007 |
|  | *Clostridium* | -0.96 | 0.25 | <0.001 | 0.01 |
|  | Lachnospiraceae | 0.89 | 0.24 | <0.001 | 0.01 |
|  | *Dorea* | -0.54 | 0.14 | <0.001 | 0.01 |
|  | *Blautia* | -0.38 | 0.11 | <0.001 | 0.02 |
|  | *Bacteroides ovatus* | 1.0 | 0.29 | 0.001 | 0.02 |
|  | *Bacteroides ovatus* | 0.75 | 0.22 | 0.001 | 0.02 |
|  | *Bacteroides* | -0.46 | 0.13 | 0.001 | 0.02 |
|  | *[Ruminococcus] torques* | -0.84 | 0.25 | 0.001 | 0.03 |
|  | Lachnospiraceae | -0.67 | 0.20 | 0.001 | 0.03 |
|  | *Dorea* | -0.60 | 0.19 | 0.002 | 0.04 |
|  | *Dorea formicigenerans* | -0.31 | 0.10 | 0.003 | 0.04 |
|  | Lachnospiraceae | -0.64 | 0.21 | 0.003 | 0.046 |
|  | Lachnospiraceae | -0.44 | 0.14 | 0.003 | 0.046 |
|  | Clostridiales | 1.1 | 0.37 | 0.003 | 0.046 |
|  | *Ruminococcus* | 0.64 | 0.21 | 0.003 | 0.046 |

SE: standard error.

**Supplementary Table 2.** ASVs with q<0.25 interaction terms between Pre and End of each treatment period as determined by MaAsLin2

| **Interaction tested** | **Treatment** | **ASV** | **Coefficient** | **SE** | ***p* value** | ***q* value** |
| --- | --- | --- | --- | --- | --- | --- |
| Fiber intake during each treatment period * time point (Pre vs. End) | RS2 | *Oscillospira* | -1.58 | 0.44 | <0.001 | 0.14 |
|  | RS4 | *⸻* | ⸻ | ⸻ | ⸻ |  |
|  | Control | *⸻* | ⸻ | ⸻ | ⸻ |  |
| Fiber intake at baseline * time point (Pre vs. End) | RS2 | Ruminococcaceae | 0.74 | 0.23 | 0.002 | 0.17 |
|  | RS4 | Clostridiales | -0.68 | 0.20 | 0.002 | 0.15 |
|  | Control | *Veillonella dispar* | -1.6 | 0.463 | <0.001 | 0.13 |
| Treatment order (Group A vs Group B) * time point (Pre vs End) | RS2 | *Blautia producta* | -0.22 | 0.071 | 0.004 | 0.21 |
|  |  | Clostridiaceae | -0.77 | 0.26 | 0.005 | 0.21 |
|  |  | Rikenellaceae | -0.36 | 0.13 | 0.006 | 0.21 |
|  |  | Ruminococcaceae | -0.66 | 0.23 | 0.005 | 0.21 |
|  | RS4 | *[Ruminococcus] gnavus* | 0.30 | 0.089 | 0.001 | 0.12 |
|  |  | *Coprococcus* | 0.32 | 0.11 | 0.005 | 0.24 |
|  | Control | *⸻* | ⸻ | ⸻ | ⸻ | ⸻ |
| *AMY1* Group (High vs Low)* time point (Pre vs End) | RS2 | *[Ruminococcus]* | 0.49 | 0.13 | <0.001 | 0.08 |
|  |  | *Erysipelotrichaceae Dielma* | 0.88 | 0.25 | 0.001 | 0.14 |
|  | RS4 | *⸻* | ⸻ | ⸻ | ⸻ | ⸻ |
|  | Control | *⸻* | ⸻ | ⸻ | ⸻ | ⸻ |
| Mean salivary amylase activity * time point (Pre vs. End) | RS2 | *Sutterella* | -0.57 | 0.16 | <0.001 | 0.13 |
|  |  | *Slackia* | -0.28 | 0.086 | 0.002 | 0.15 |
|  |  | *Methanobrevibacter* | 0.22 | 0.073 | 0.003 | 0.24 |
|  | RS4 | *Lachnospiraceae* | -0.58 | 0.15 | <0.001 | 0.19 |
|  | Control | *Sutterella* | 0.62 | 0.14 | <0.001 | 0.045 |

ASVs in which we observed interaction effects (q<0.25) between candidate predictors (energy-adjusted dietary fiber intake during treatment, energy-adjusted dietary fiber intake during baseline, treatment order, *AMY1* Group, and mean SAA) and time point type (Pre vs End of treatment). Since this was a screening step we raised our threshold of significance to q<0.25. SE: Standard Error.

**Supplementary Table 3.** Study cracker formulations

| Ingredient | Control | RS2 | RS4 |
| --- | --- | --- | --- |
| Bread flour | 49.85% | 29.40% | 45.27% |
| Amioca TF starch | 24.72% | - | - |
| HiMaize 260 | - | 44.78% | - |
| VersaFibe 1490 | - | - | 29.95% |
| Unsalted Butter | 17.89% | 17.59% | 18.05% |
| Sugar | 2.41% | 2.36% | 2.44% |
| Salt | 1.15% | 1.12% | 1.15% |
| Water | 3.99% | 4.76% | 3.14% |
| TOTAL | 100.0% | 100.0% | 100.0% |

RS2 crackers contained HI-MAIZE® 260 starch (56% RS), RS4 crackers contained VERSAFIBE^TM^ 1490 starch (85% RS), and Control crackers contained AMIOCA™ TF starch (100% digestible starch) (Ingredion Inc., Bridgewater, N.J., USA).

**Supplementary Table 4.** RS and total dietary fiber content in study crackers

|  | Control Cracker | RS2 Cracker | RS4 Cracker |
| --- | --- | --- | --- |
| RS | < 2.4 g | 21.27 g | <2.4 g |
| Total dietary fiber | 1.92 g | 35.64 g | 36 g |

Content of resistant starch and total dietary fiber in the study crackers as analyzed by Medallion Laboratories (Minneapolis, MN) using the methods AOAC 2002.02 for RS and AOAC 991.43 for dietary fiber. AOAC 2002.02 method is known to underestimate the amount of RS4 and therefore that measurement should be interpreted with caution (1). Amounts in the table are per 120 g of crackers, which was the daily portion of study crackers per each participant after the gradual dose escalation period.
